# P0: Progress to zero: A simple metric to measure COVID-19 progress by country/region

**DOI:** 10.1101/2020.05.21.20109298

**Authors:** Juan M. Lavista Ferres, Ruth B Etzioni, William B Weeks

## Abstract

The current COVID-19 pandemic is having a devastating impact on the world. As of May 8th, 2020, more than half of the world is in lockdown, 3.6 million people have been infected, and over 250,000 of them have died. This is the first pandemic in human history that has been tracked on a daily basis across the world. Currently, progress is being tracked by a series of metrics, including new cases, deaths, and the effective reproduction number over time (R(t)), which epidemiologists have emphasized.

While it is an important measure, R(t) is not observable and is dependent on the absolute number of active cases; therefore, two geographies could have a similar R(t) but be having very different experiences. Here, we propose a metric to monitor progress on addressing the COVID pandemic that we name “progress to zero” or P0. P0 corresponds to the percentage decline from a previously recorded peak level. The metric ranges from 0% (representing a geography that has not yet peaked) to 100% (representing a geography wherein 0 cases have been recorded for at least seven days). Our metric helps leaders focus on progress to a desired goal, easily can be tracked over time, can quickly identify retreat from progress, and is comparable across geographies regardless of their size.

## Introduction

Objectively evaluating progress on addressing the COVID-19 pandemic will require multiple metrics, including cases, deaths, testing capacity, and hospitalizations over time. [1]. One metric that epidemiologists focus on is the change in effective reproductive number over time (R(t)) [2].

Epidemiologists argue that tracking R(t) is a way to manage the COVID-19 crisis [3–6]. R(t) conveys whether the number of cases is growing (R(t)>1), has plateaued (R(t)~1), or is declining (R(t)<1).

However, R(t) is a blunt metric that misses nuance. R(t) is interpretable, but it is not observable: it must be estimated from the available data on confirmed cases over time. Further, because different geographies have different individual, biological, and environmental factors, R(t) may not be comparable across different geographic settings.

For example, on May 5th 2020, the R(t) for the United States (US), the United Kingdom (UK), and Australia were all near 1, suggesting that the number of cases had plateaued. [1] Nevertheless, the lived experience of the three countries on May 8th was very different [7] (Figure 1). While US and UK cases had plateaued near the peak number of cases but continued with large absolute numbers of new cases, Australia, had fewer than 20 new cases per day and had several 0 death days [8]. Similarly, while R(t) was similar in Hungary and Argentina on May 8th, the trajectory of new cases was very different (Figure 2).

**Figure 1.**
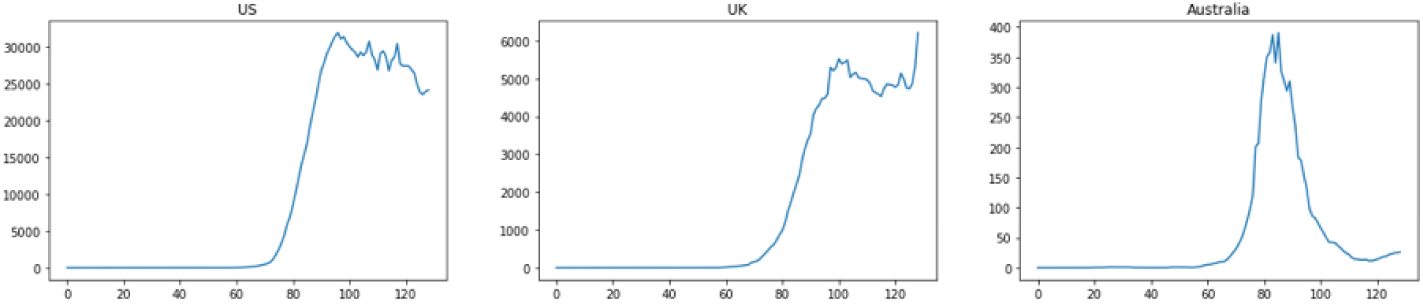
New COVID-19 cases per day, US, UK, Australia [5]

**Figure 2.**
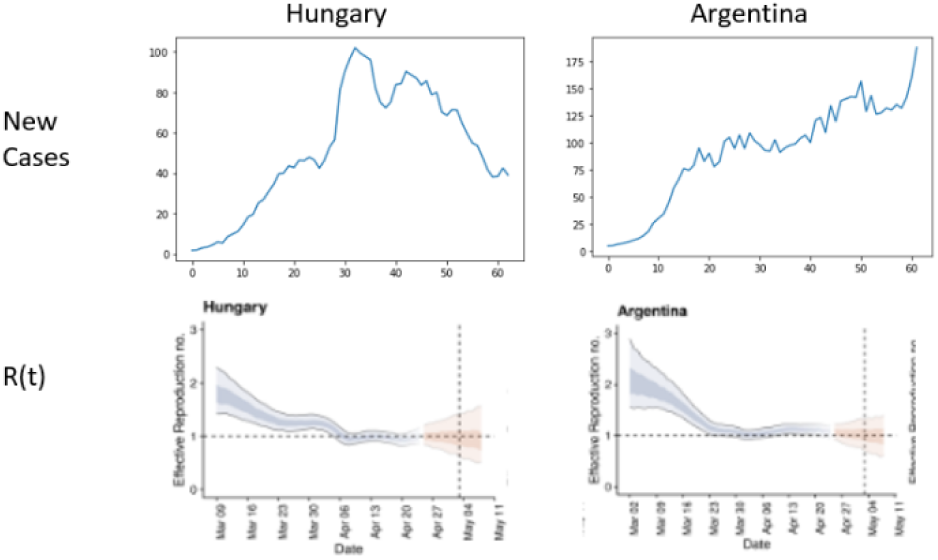
New COVID-19 cases per day [5], R(t) [1] for Hungary and Argentina

Clearly, the relevance of an R>1 depends on the absolute number of active cases. In the absence of knowing the absolute numbers of cases, it is difficult to derive conclusions from an R(t) graphic.

## Methods

Here, we propose an additional metric for leaders to measure success in addressing the COVID-19 crisis: progress to zero (P0). This is defined as the percentage decline from a previously recorded peak level.

P0 at time i is defined one minus the ratio of A(i) to B(i), where A is the 7-day moving average of new cases ending at day i and B is the 7-day moving average of new cases corresponding to the historic peak. While the measure could be defined using a shorter moving average interval or using a weighted moving average, we use a simple 7-day moving average to illustrate its calculation and use.

Because the measure would not be stable with small numbers of cases, we recommend limiting measure application to geographies with at least 100 COVID-19 cases confirmed within a region.

Defining a 7-day moving average of cases and i as the current day, the calculations are provided below:

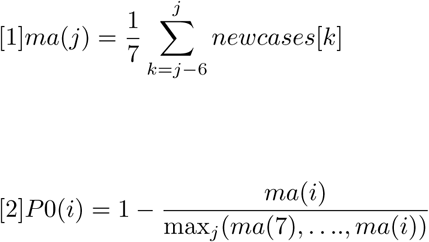

## Results

As an example, applying this metric on May 8th, 2020, demonstrates considerable progress disparity across Australia, the US, the UK, Hungary, and Argentina (Table 1).

**Table 1.** Progress to zero cases and R(t) [1]

| | Progress | $R(t)$ |
| --- | --- | --- |
| **Australia** | 93% | 1 (0.5 – 1.4) |
| **United_Kingdom** | 0% | 1 (0.9 – 1) |
| **United_States_of_America** | 24% | 1(1 – 1.1) |
| **Hungary** | 62% | 1(0.9 -1.2) |
| **Argentina** | 0% | 1.1(1 – 1.2) |

Effectively, the measure suggests that there is no progress if the number of cases has not peaked, 100% progress if there are zero cases for at least one week, and intermediate progress (which is comparable across geographies) if the number of cases is declining. Particularly in anticipation of the ongoing growth of viral spread and multiple potential waves, it is important to recognize that a country could peak several times. With each new peak, the metric will reset, with the global peak being the new baseline (Figure 3 gives an example).

**Figure 3.**
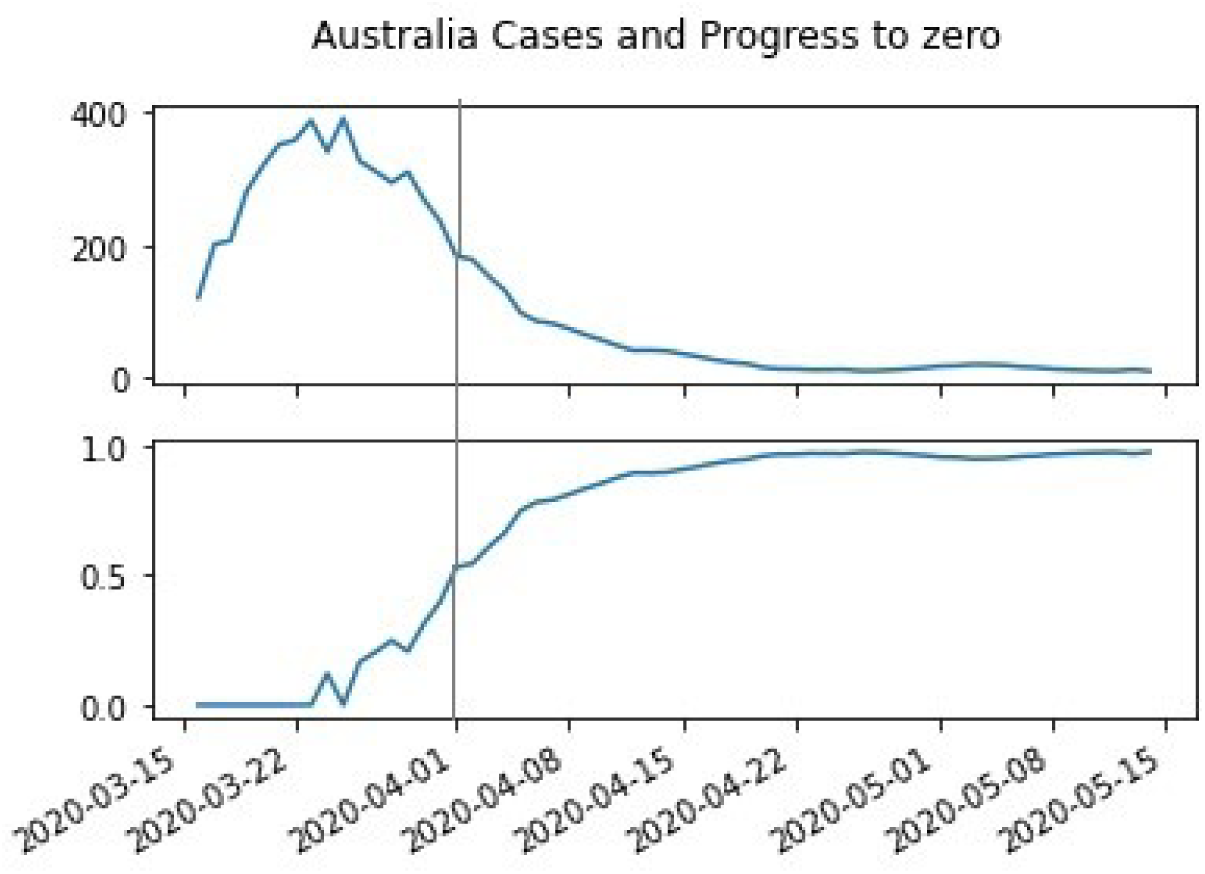
Australia new COVID-19 [5] cases and progress to zero (P0)

The measure also captures the importance of controlling the growth of cases. As of April 1, 2020, Australia had 200 new cases per day, similar to that of Argentina had as of May 1st. However, progress for Australia was 50%, while Argentina’s progress was zero. Independent of the number of new cases that a country has, as long as the country cannot reduce them, cases will continue to grow, making future control more challenging. On the other hand, if a country has learned how to reduce the development of new cases, absent policy changes, control of case spread should continue.

### Measuring Deaths, cases, and hospitalizations

Using data from the European Center for Disease Control, [4] as of May 8, 2020, only 5 countries had made at least 95% progress towards 0 cases using our metric (China, South Korea, Thailand, Israel, and Switzerland) (Figure 5). Disconcertingly, 30 countries have made less than 5% progress and may not have reached their global peak yet.

**Figure 4.**
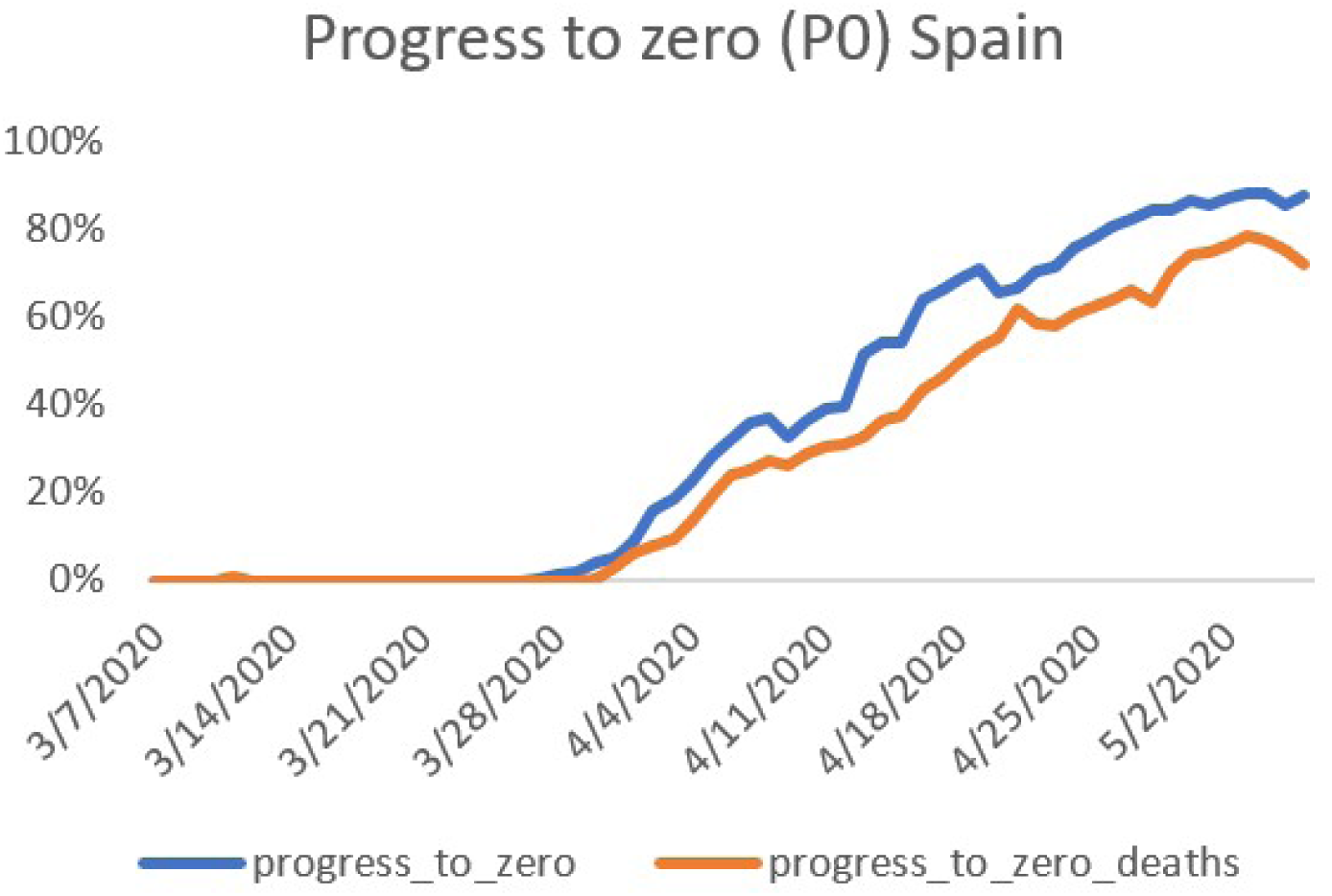
Progress to zero (P0) cases and deaths in Spain

**Figure 5.**
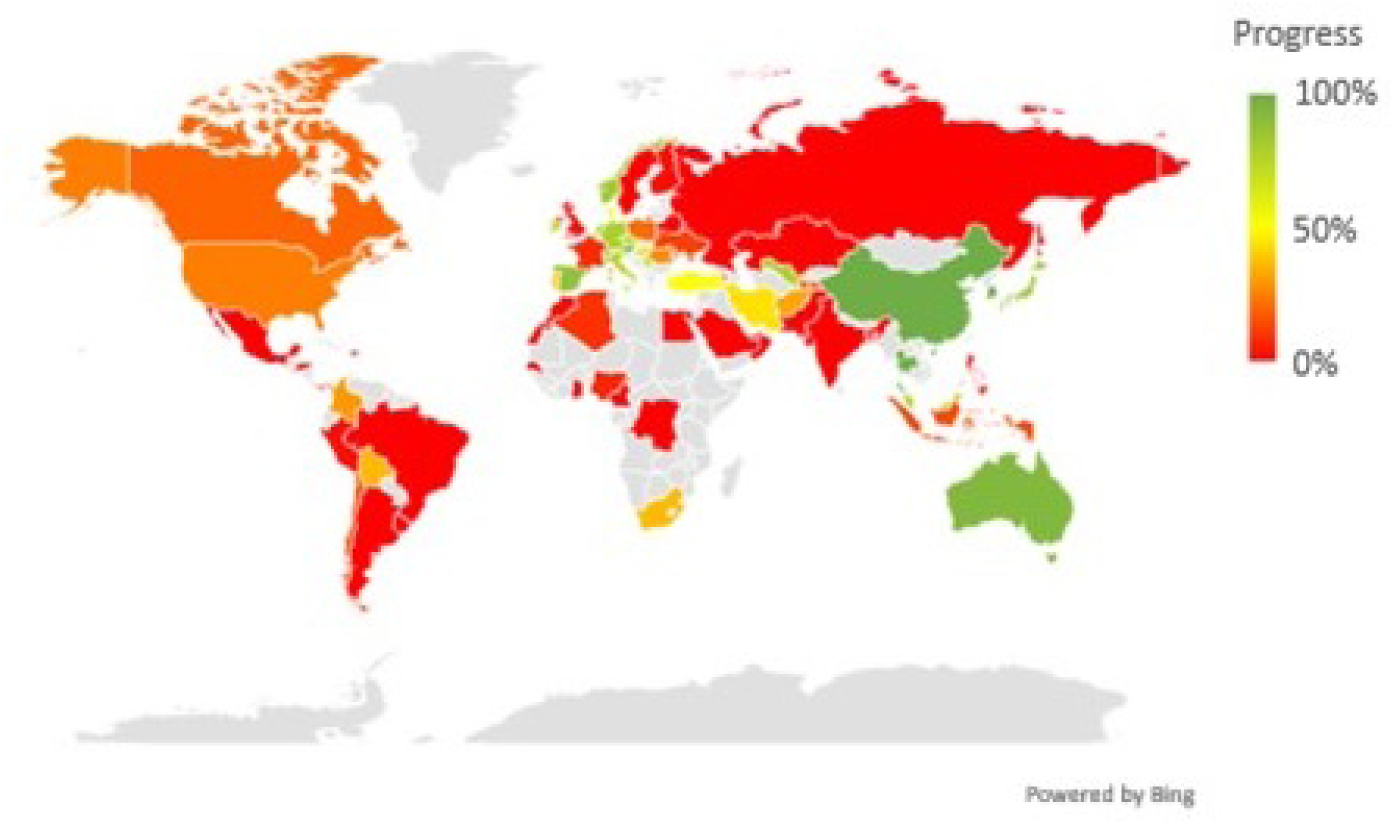
Progress to zero cases as of May 8th, 2020

## Data Availability

All data is publicly available. 
the code to compute the metric is available here
https://github.com/jlavista/progresstozero/blob/master/Progress%20to%20zero%20P0.ipynb

